# Misalignment between circadian preference and accelerometer-derived actual sleep-wake cycle is associated with increased risk of cardiometabolic diseases: a prospective cohort study in UK Biobank

**DOI:** 10.1101/2024.06.28.24309628

**Authors:** Yue Chen, Tingting Geng, Xinming Xu, Zhicheng Zhang, Lili Huang, Haiyang Dong, Huan Yu, Xiang Gao, Liang Sun

## Abstract

**Background:** Evidence has linked several circadian disruption indicators, such as social jetlag and shift work, to adverse health outcomes, however, associations of misalignment between circadian preference and actual sleep-wake cycle with cardiometabolic diseases (CMDs) remains unclear. We aimed to prospectively investigate the associations of the circadian misalignment with CMDs including type 2 diabetes (T2D), chronic heart diseases (CHD), and stroke, and to uncover potential mechanisms linking circadian misalignment and CMDs.

**Methods:** A total of 60,965 participants from the UK Biobank study without baseline CMDs and followed-up for an average of 7.9 years were included in the current analysis. Circadian misalignment was defined as discrepancies between self-reported chronotype and accelerometer-derived midpoint of sleep to detect its association with CMDs. Incident CMDs were derived from primary care, hospital inpatient, death registry, and self-reported source of data. Cox proportional hazards regression models were used to compute the hazard ratio (HR) and confidence intervals (CIs) on the association between circadian misalignment (quintiles of residuals of midsleep on chronotype) and incident CMDs.

**Results:** U-shaped associations were found of the circadian misalignment with incident T2D and CHD after adjusting the potential confounders. Compared to individuals with aligned midsleep and circadian preferences (Q3), those with advanced and delayed circadian misalignment had higher risks of T2D [HR (95%CI) 1.22 (1.03, 1.45) in Q1 and 1.39 (1.18, 1.62) in Q5]. However, only delayed circadian misalignment was significantly associated with an increased risk of CHD [HR (95%CI) 1.15 (1.01, 1.31) in Q4 and 1.16 (1.02, 1.33) in Q5]. Liver function, lipid and glucose metabolism, and inflammatory markers partially explained the observed circadian misalignment and CMDs association (mediation proportion 12.3-44.6% for T2D, 8.8-20.5% for CHD). Moreover, the association between delayed circadian misalignment and CMDs was more prominent in women (for T2D, *P_interaction_*=0.03) and in younger adults (for CHD, *P_interaction_*=0.02) compared to their counterparts. Additionally, early chronotype [HR (95%CI): 1.19 (1.06, 1.34)] rather than late chronotype was associated with an increased risk of incident T2D.

**Conclusion:** Both advanced and delayed circadian misalignment were associated with increased risks of CMDs, suggesting potential benefits of aligning actual sleep-wake cycles with individual circadian preferences.

**Novelty and Significance:** *What is known?:*

- Cardiometabolic diseases such as type 2 diabetes, coronary heart disease, and cerebrovascular diseases have become major concerns in global public health over recent decades.
- Evidence has linked several circadian disruption indicators to adverse health outcomes: People living with late chronotype (circadian preference), late sleep timing, long social jetlag (the difference between midsleep on work-free days and work days), and shift work were prone to introduce circadian disruptions and had higher risks of cardiometabolic diseases.
- Little is known about whether and how the misalignment between circadian preference and actual sleep-wake cycle is associated with cardiometabolic diseases, particularly in large prospective cohort studies.

*What New Information Does This Article Contribute?:*

- This is the first study to propose a new metrics (misalignment between circadian preference and actual sleep-wake cycle) to shed some lights on quantifying human circadian disruption in general population of differed work schedule and in population with less disrupted circadian rhythm.
- Both advanced and delayed sleep-wake cycle comparing to individual circadian preference were associated with increased risks of cardiometabolic diseases. Population may benefit from keeping an actual sleep-wake cycle in accordance with their circadian preference.
- Early chronotype rather than late chronotype were found to associate with an increased risk of incident T2D, independent of actual sleep-wake cycle.
- Age and sex modified the associations between circadian misalignment and cardiometabolic diseases, while liver function, lipid and glucose metabolism, and inflammatory markers partially mediated the associations.

## Introduction

Cardiometabolic diseases (CMDs) such as type 2 diabetes (T2D), coronary heart disease (CHD), and cerebrovascular diseases have become major concerns in global public health over recent decades^1^. The multifactorial nature of the etiology of these diseases involves a complex interplay of genetic, lifestyle, and environmental factors, presenting great challenges on the prevention and prognosis of CMDs^2^. Therefore, more efforts are needed to develop potential strategies to combat this escalating phenomenon.

Circadian disruption indicated a wide range of misalignment upon individual internal-internal timing as well as internal-external rhythm^3^. In field-based studies, metrics for circadian disruption mainly rely on rest-activity measurement and disrupted sleep behavior, e.g., social jetlag and shift work^3^. Epidemiological evidence suggested that people living with late chronotype (circadian preference), late sleep timing, long social jetlag (the difference between midsleep on work-free days and work days), and shift work were prone to introduce circadian disruptions^4^ and had higher risks of CMDs^5-9^. However, current widely used proxies for circadian disruption (such as social jetlag and shift work) either largely require regular work schedule, or lack of generalizability to the general population who works non-shift and less deviated from internal clock^10^. In addition, the majority of prior epidemiological studies on circadian disruptions and CMDs were cross-sectional designs^6, 11, 12^, only very limited prospective analysis was conducted in large population^13^. Furthermore, advancements in sensor element technology and accelerometer now allow the objective measurement of human sleep-wake cycle, physical activity, and ambient environmental zeitgebers. Unlike biomarkers that objectively measured human circadian rhythm requiring repeated or invasive procedures^14, 15^, accelerometers enable a non-intrusive way to consistent tracking rest-activity cycles^14-16^. Notably, over 80% population in a large database (the Munich ChronotType Questionnaire database) reported using an alarm clock and accumulated sleep debts during working week^17^, indicating a large proportion of population suffer from misalignment between circadian preference and their actual sleep-wake cycle. However, little is known about whether and how the misalignment between circadian preference and actual sleep-wake cycle is associated with CMDs, particularly in large prospective cohort studies.

Thus, based on a large-scale cohort study, the UK Biobank, we used the information on accelerometer-derived sleep-wake cycle, the self-reported circadian preference, and comprehensive diseases diagnosis resources 1) to prospectively analyze the association of circadian misalignment with CMDs such as T2D, CHD, and stroke, and 2) to reveal the potential mechanism underlying the relationship between circadian misalignment and CMDs, through mediation analysis by plasma biomarkers and anthropometric indices.

## Methods

### Study design and participants

The UK Biobank, initiated in 2006-2010, is a large-scale prospective cohort study containing comprehensive health information from over half a million adult UK participants. The UK Biobank was granted ethical approval from the North West Multi-center Research Ethical Committee (reference # 11/NW/0382) and research is performed in accordance with the Declaration of Helsinki. This research has been conducted using the UK Biobank Resource under application number 96083.

Among 502,505 participants at baseline in the UK Biobank, 74,123 participants with complete and valid information on both baseline sleep questionnaire and accelerometer derived information were included, after further excluding 7,052 participants with prevalent diabetes, stroke, or CHD at the time of monitoring activity, 389 participants with baseline random glucose level ≥ 11.1 mmol/L or HbA1c ≥ 48 mmol/mol, 5,418 participants reported jobs involves shift work, and 299 participants with measured sleep duration < 4 hours or > 12 hours, 60,965 participants were included in final analysis (**Figure 1**).

**Figure 1.**
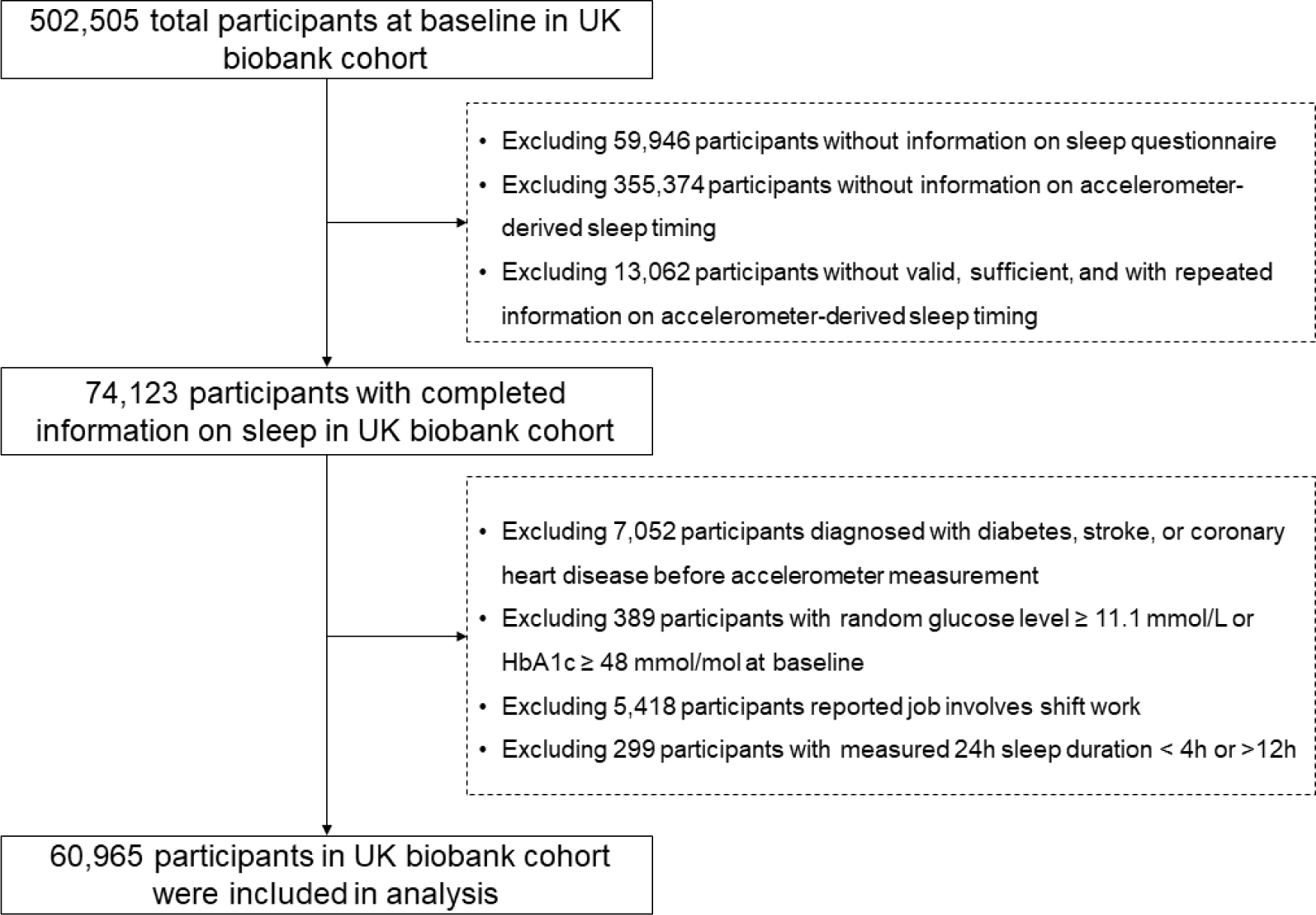
Flow chart of the study.

### Sleep assessment

Sleep status was evaluated through both subjective touchscreen sleep questionnaires and objective actigraphy monitors.

#### Sleep questionnaire

At baseline recruitment, information on personal chronotype, 24h sleep duration, snoring, insomnia, and daytime sleepiness was collected by a self-administrated touchscreen sleep questionnaire. They were collected by the following questions: “Do you consider yourself to be? (definitely a ‘morning’ person, more a ‘morning’ person than an ‘evening’ person, more an ‘evening’ person than a ‘morning’ person, definitely an ‘evening’ person, do not know, prefer not to answer)”, “About how many hours sleep do you get in every 24 hours? (please include naps)”, “Does your partner or a close relative or friend complain about your snoring? (yes, no, do not know, prefer not to answer)”, “Do you have trouble falling asleep at night or do you wake up in the middle of the night? (never/rarely, sometimes, usually, prefer not to answer)”,” How likely are you to doze off or fall asleep during the daytime when you don’t mean to? (e.g., when working, reading or driving) (never/rarely, sometimes, often, do not know, prefer not to answer, all of the time)”. For all the above questions, answers with “do not know”, “prefer not to answer” were considered as missing. Healthy sleep quality score was calculated from questionnaire derived chronotype, insomnia, snore, daytime sleepiness, and sleep duration, see **Supplementary TableS1.**

#### Actigraphy

During June 2013 and January 2016, physical activity and sleep measurements were recorded via a wrist-worn accelerometer (Axivity AX3), which is a commercial version of the Open Movement AX3 open-source sensor (https://github.com/digitalinteraction/openmovement) designed by Open Lab, Newcastle University. Exquisite processes on data collection and curation have been described previously^18^. Briefly, participants were instructed to wear the accelerometer upon receipt and proceed with their usual routines, affixing it to their dominant wrist. They were notified that the device would activate automatically upon delivery and deactivate after a week. At the end of this period, participants returned the device to the central office using a provided pre-paid envelope. R package GGIR was used to clearing, calibrating, and deriving information on timing of sleep, moderate to vigorous physical activity, and light exposure^19, 20^.

### Ascertainment of circadian misalignment

The morning-evening preference, chronotype, was collected by one validated question from the Horne and Östberg Morningness-Eveningness Questionnaire (MEQ)^21^. This question indicated a good correlation with the overall score of the MEQ (r = 0.72)^22^, and was also employed in other large cohort studies to indicate individual circadian preference^13, 23^. Additionally, information on participants timing of onset and end of sleep was collected by wearing an actigraphy on dominant wrist through seven consecutive days and derived by R package GGIR. Nighttime sleep duration was calculated as the difference between time point of sleep end and sleep onset. Midsleep of weekdays, weekend, and weekly average can be calculated using the following formula, using the average value of sleep onset and sleep duration on weekdays, weekend, and the whole week, respectively.

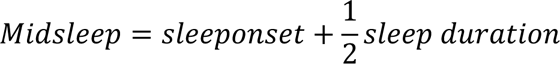

The misalignment between circadian preference (chronotype) and midsleep was calculated as the residuals of midsleep on chronotype or the difference between midsleep and chronotype (subtracting group numbers of chronotype from midsleep)^24^, detailed calculation and categorization can be found in **Supplementary Method**.

### Cardiometabolic outcomes

Newly onset of T2D, CHD, and stroke (including ischemic stroke and hemorrhage stroke subtypes) were individually evaluated and collectively considered when calculating CMD incidence. Data on data showcase-first occurrence (Category 1712) were used to derive the first occurrence date of diseases. Two main classification systems of clinical coding were used in the linked health data: International Classification of Diseases (ICD) and Read. Information on primary cares (Read v2 and Read CTV3), hospital inpatient (ICD10 and ICD9), death register (ICD10), cancer register (ICD10 and ICD9), and self-reported medical condition were gathered and mapped to 3-character ICD-10 to ascertain the source and date of the first occurrence of diseases. Detailed method can be found here (https://biobank.ndph.ox.ac.uk/showcase/ukb/docs/first_occurrences_outcomes.pdf). ICD code and field ID of outcomes were described in **Supplementary table S2**. Incident cases were defined as cases with a later date of first occurrence in comparison with the date of measurement of baseline information and accelerometer.

### Covariates

Baseline information on demographic and socioeconomic status (age, sex, education, Townsend deprivation index, region, ethnic group, employment status) and family history of diseases were collected in assessment center when participants were initially recruited. Lifestyle factors including drinking and smoking status, tea and coffee intake, as well as a detailed 29-item food frequency questionnaire on dietary intakes were also evaluated in assessment center by touchscreen questionnaire. Diet intake information was converted to a Mediterranean diet score to indicate dietary quality^25^.

Additionally, baseline body weight, height, waist circumference, hip circumference, and blood pressure were measured in assessment center under standard process. Body mass index was calculated as weight (kg) divided by height (m) squares, and waist-hip-ratio was also calculated. Blood samples were collected at recruitment, separated by components and stored at UK Biobank (−80°C and LN2). Sample collection and processing has been previously described in detail (https://biobank.ndph.ox.ac.uk/showcase/ukb/docs/biomarker_issues.pdf). A range of key biochemistry markers (hepatic and renal function, inflammatory cytokines, lipids and glucose metabolism) were used in the current analysis. Levels of glucose and HbA1c at baseline were employed to identify participants with prevalent diabetes. Levels of Low-Density Lipoprotein was also used in analysis, as it was closely related to cardiometabolic outcomes^26^.

### Statistical analysis

All statistical analyses were performed using SAS 9.4 (SAS Institute Inc) and R version 3.4.2. Two-sided *P*<0.05 were regarded as the level of significance. Descriptive data of participants at baseline were shown as number (%) for categorical variables and mean (standard deviation) for continuous variables. See **Supplementary Method** for the adjusted circadian misalignment according to baseline characteristics and selection of covariates in the following models.

Cox proportional hazard models were used to investigate the associations between circadian misalignment (quintiles of residuals of midsleep on chronotype) and incident cardiometabolic events. All cox models have passed the proportional hazards assumption by Schoenfield Residuals Test. Model 1 was adjusted for age (tertiles), sex (men/women), and ethnic group (White British/Non-White British), and Model 2 was additionally adjusted for region (urban/town/village), Townsend deprivation index (tertiles), employment (in-paid employment/retired/without fixed employment), qualification (college or university degree/A levels or equivalent/O levels or GCSEs or equivalent/none of the above), and ambient noise level (tertiles). Model 3 (main model) was further adjusted for smoking status (never/ever/current), drinking status (never/ever/current), tea consumption (<2/2-4/≥5 cups), coffee consumption (0/0-2/≥3 cups), accelerometer-derived moderate to vigorous physic activity (tertiles), and healthy sleep quality score (<2/2-3/>3). All continuous covariates were converted to tertiles to clearer show their relationship with circadian misalignment (Figure S1) and CMDs. Since missing values for all covariates were less than 6% (5.9% for healthy sleep quality score, and 1.2% for ambient noise level, and less than 0.2% for other covariates), they were imputed as the mode^12^. Plots on restricted cubic splines (four knots on 0.05, 0.35, 0.65, and 0.95, respectively) and *P* for nonlinear (ANOVA test^27^) were used to show the nonlinear relationship between circadian misalignment and cardiometabolic events. Moreover, risk of cardiometabolic events were also evaluated by circadian misalignment using difference between midsleep and chronotype (advanced/intermediate/delayed group), as well as chronotype and midsleep individually. When investigating the association of midsleep and chronotype with CMDs, they were further mutually adjusted.

Sensitivity analyses were conducted by 1) recategorizing the main exposure, circadian misalignment (residuals of midsleep on chronotype), into three groups [advanced (Q1), intermediate (Q2+Q3+Q4), delayed (Q5) group]; 2) recalculating the main exposure, circadian misalignment, as the absolute value of the residuals of midsleep on chronotype, and categorized it into tertiles. The linear trend of HRs over tertiles was assessed by *χ^2^* test using these tertile numbers as continuous variables; 3) based on main model (Model 3), additionally adjusted for family history of diabetes (when outcome is T2D), family history of cardiovascular diseases (when outcome is stroke and CHD), or family history of diabetes and cardiovascular diseases (when outcome is CMDs), and time span between baseline recruitment and accelerometer assessment, as they were suggested confounders in previous studies. Age was adjusted in the form of continuous variable to reduce potential residual confounding; 4) Based on main model, further adjusted for BMI, LDL, and SBP, as they may act both as underlying mediators and confounders; 5) excluding upper and lower 0.5% value of residuals of midsleep on circadian preference; 6) calculating propensity score to balance the distribution of baseline characteristics among differed circadian misalignment group. Propensity score weights for multiple treatments and average treatment effect on the treated was calculated by the R package Twang and using the method of gradient boosting machine^29, 30^; 7) using ICD-10 code E11 instead of pooling E11 and E14 together to define T2D; and 8) including participants who worked with shift work in analysis, as shift work was an important contributor of circadian misalignment.

In secondary analysis, causal mediation analyses were conducted by R package CMAverse^28^. Average total effect hazard ratio, nature direct hazard ratio, nature indirect hazard ratio, and proportion mediated by potential mediators were shown. Mediators were log-transformed and standardized to show HRs (95% CI) per standard deviation. Proportional hazards assumptions were also confirmed. See **Supplementary Method** for the selection of potential mediators. Furthermore, subgroup analyses were conducted by age (<57 /≥57 years (median)), sex (women/men), ethnic (White British/non-White British), residence (Urban/town or village), socioeconomic status (<median/≥median), and healthy sleep quality score (<4/≥4). The multiplicative interaction between characteristics and circadian misalignment was also detect by additionally adding an interaction term to the model.

## Results

### Baseline characteristics and risk factors of delayed circadian misalignment

60,965 participants aged 39 to 70 years at baseline were included in analysis. Compared to advanced circadian misalignment group, participants of delayed circadian misalignment group were prone to be older, women, and non-White British. They were more likely to live in urban areas and be exposed to higher ambient noise level, have lower education levels and socioeconomic status, and work without pay. In terms of lifestyle, participants predisposed to delayed circadian misalignment tended to be current smokers and drinkers, engage in less moderate to vigorous physical activity, drink more tea and less coffee, and have worse sleep quality **(Table 1 and Figure S1)**.

**Table 1.** Baseline characteristics of 60965 study participants from UK Biobank according to circadian misalignment^a^.

|  | Circadian misalignment<sup>b</sup> |  |  |
| --- | --- | --- | --- |
|  | Advanced group<br>(N=12358) | Intermediate<br>group<br>(N=36464) | Delayed group<br>(N=12143) |
| **Basic information and lifestyle factors** |  |  |  |
| **Women (%)** | 7088 (57.4) | 22449 (61.6) | 7466 (61.5) |
| **Urban area (%)** | 10141 (82.9) | 30038 (83.1) | 10283 (85.6) |
| **Age at baseline (years)** | 54.4 (8.2) | 56.1 (7.8) | 57.5 (7.2) |
| **Townsend deprivation index<sup>c</sup>** | -1.7 (2.8) | -1.9 (2.7) | -1.5 (2.9) |
| **Level of education (%)** |  |  |  |
| College or University degree | 5522 (44.7) | 17084 (46.9) | 5231 (43.1) |
| A levels/AS levels or Equivalent | 1674 (13.6) | 4837 (13.3) | 1595 (13.1) |
| O levels/GCSEs or Equivalent | 2504 (20.3) | 7343 (20.1) | 2448 (20.2) |
| None of the above | 2658 (21.5) | 7200 (19.8) | 2869 (23.6) |
| **White British (%)** | 11956 (96.8) | 35551 (97.5) | 11667 (96.1) |
| **Family history of diabetes (%)** | 2367 (19.2) | 7019 (19.3) | 2358 (19.4) |
| **Family history of cardiovascular diseases (%)** | 6726 (54.4) | 20796 (57.0) | 7196 (59.3) |
| **On-paid work (%)** | 8331 (67.4) | 22289 (61.1) | 6361 (52.4) |
| **Drinking status (%)** |  |  |  |
| Never drinker | 372 (3.0) | 952 (2.6) | 371 (3.1) |
| Ever drinker | 365 (3.0) | 817 (2.2) | 382 (3.2) |
| Current drinker | 11621 (94.0) | 34695 (95.2) | 11390 (93.8) |
| **Smoking status (%)** |  |  |  |
| Never smoker | 7212 (58.4) | 21802 (59.8) | 6627 (54.6) |
| Ever smoker | 4302 (34.8) | 12619 (34.6) | 4427 (36.5) |
| Current smoker | 844 (6.8) | 2043 (5.6) | 1089 (9.0) |
| **Daily average coffee intake (cups)** | 2.0 (2.0) | 1.9 (1.9) | 2.0 (2.0) |
| **Daily average tea intake (cups)** | 3.2 (2.7) | 3.3 (2.6) | 3.4 (2.8) |
| **Mediterranean dietary score (points)** | 2.9 (1.4) | 2.9 (1.4) | 2.9 (1.4) |
| **Daily average time on moderate to vigorous physical activity per day<sup>d</sup> (min)** | 117.0 (53.2) | 115.4 (50.5) | 104.9 (50.1) |
| **Daily average ambivalent Light exposure<sup>e</sup> (lux)** | 781.2 (161.6) | 803.7 (149.0) | 788.3 (154.7) |
| **Daily average ambivalent noise<sup>f</sup> (dB)** | 51.2 (4.2) | 51.1 (4.1) | 51.2 (4.3) |
| **Circadian and sleep** |  |  |  |
| **Chronotype** |  |  |  |
| Early | 2960 (24.0) | 9262 (25.4) | 3449 (28.4) |
| Intermediate early | 4702 (38.1) | 14507 (39.8) | 3936 (32.4) |
| Intermediate late | 3485 (28.2) | 9944 (27.3) | 3334 (27.5) |
| Late | 1211 (9.8) | 2751 (7.5) | 1424 (11.7) |
| **Weekly average midsleep<sup>g</sup> (hh:mm)** | 02:06 (00:48) | 03:18 (00:30) | 04:24 (00:48) |
| **Midsleep on weekdays (hh:mm)** | 02:06 (00:54) | 03:18 (00:30) | 04:36 (00:54) |
| **Midsleep on weekends (hh:mm)** | 02:12 (01:24) | 03:24 (00:48) | 04:42 (01:06) |
| **Social jetlag<sup>h</sup> (hours)** | 0.1 (1.6) | 0.2 (0.8) | 0.1 (1.2) |
| **Residuals of midsleep on chronotype<sup>b</sup>** | -1.2 (0.7) | 0.0 (0.4) | 1.3 (0.6) |
| **Healthy sleep quality score (points)<sup>i</sup>** | 3.2 (0.9) | 3.3 (0.9) | 3.1 (0.9) |

<sup>a</sup> Values are mean and standard deviation or number and percentage.
<sup>b</sup> Circadian misalignment was calculated by the residuals of midsleep on chronotype, while the negative or positive value of circadian misalignment indicated an advanced or delayed real sleep-wake cycle comparing to circadian preference. The first quintile of residuals of midsleep on chronotype was defined as the advanced group, demonstrating an advanced sleep-wake cycle relative to their circadian preference. Conversely, the fifth quintile was defined as the delayed group, which indicated a delayed sleep-wake cycle in comparison to circadian preference. The remaining quintiles (Q2, Q3, Q4) combined together were defined as the intermediate group.
<sup>c</sup> Townsend deprivation index is calculated based on the preceding national census output areas. Each participant is assigned a score corresponding to the output area in which their postcode is located. Higher value indicated a poorer socioeconomic status.
<sup>d</sup> Accelerometer-derived time on moderate or vigorous activity per day, weighted by weekends and weekdays based on R package GGIR<sup>19</sup>.
<sup>e</sup> Accelerometer-derived mean waking hours' light exposure per day based on R package GGIR<sup>19</sup>.
<sup>f</sup> Day-evening-night equivalent noise level: A-weighted local environment annual average noise level measured over the 24-hour period.
<sup>g</sup> Accelerometer-derived time on average midpoint between sleep onset and sleep end, derived and weighted by weekends and weekdays based on R package GGIR<sup>19</sup>.
<sup>h</sup> Difference between accelerometer-derived time of midsleep on weekends and workdays, derived and weighted by weekends and weekdays based on R package GGIR<sup>19</sup>.
<sup>i</sup> The healthy sleep quality score is calculated by the sum of five components (chronotype, 24-hour sleep duration, insomnia, snoring, and daytime sleepiness) from a touchscreen sleep questionnaire. It ranges from 0-5, with a higher score indicating a better sleep quality.

### Association between circadian misalignment and incident cardiometabolic events

During an average follow-up of 7.86 years, 1,526 (2.50%), 684 (1.12%), and 2,186 (3.59%) participants developed incident T2D, stroke, and CHD, respectively, yielding an overall 4,084 (6.70%) participants developing CMDs in total.

After fully adjustment in main model (Model 3), compared to individuals with matched midsleep and circadian preferences (Q3), both the advanced and delayed circadian misalignment were associated with higher type 2 diabetes [HR (95%CI) 1.22 (1.03, 1.45) in Q1 and 1.39 (1.18, 1.62) in Q5] and CMDs [HR (95%CI) 1.12 (1.02, 1.25) in Q1 and 1.21 (1.10, 1.33) in Q5]. However, only delayed circadian misalignment (Q4 and Q5) was significantly related to increased risk of CHD [HR (95%CI) 1.15 (1.01, 1.31) in Q4 and 1.16 (1.02, 1.33) in Q5] **(Table 2)**. Circadian misalignment was not associated with incidence of stroke or stroke subtypes **(Table 2** and **Table S3)**. When calculating circadian misalignment by the difference between midsleep and chronotype, participants with delayed circadian misalignment had higher risk of developing T2D [HR (95%CI): 1.19 (1.06, 1.33)] and CMD [HR (95%CI): 1.10 (1.03, 1.17)]. Consistently, neither type of circadian misalignment was associated with incidence of stroke **(Figure 2)**.

**Figure 2.**
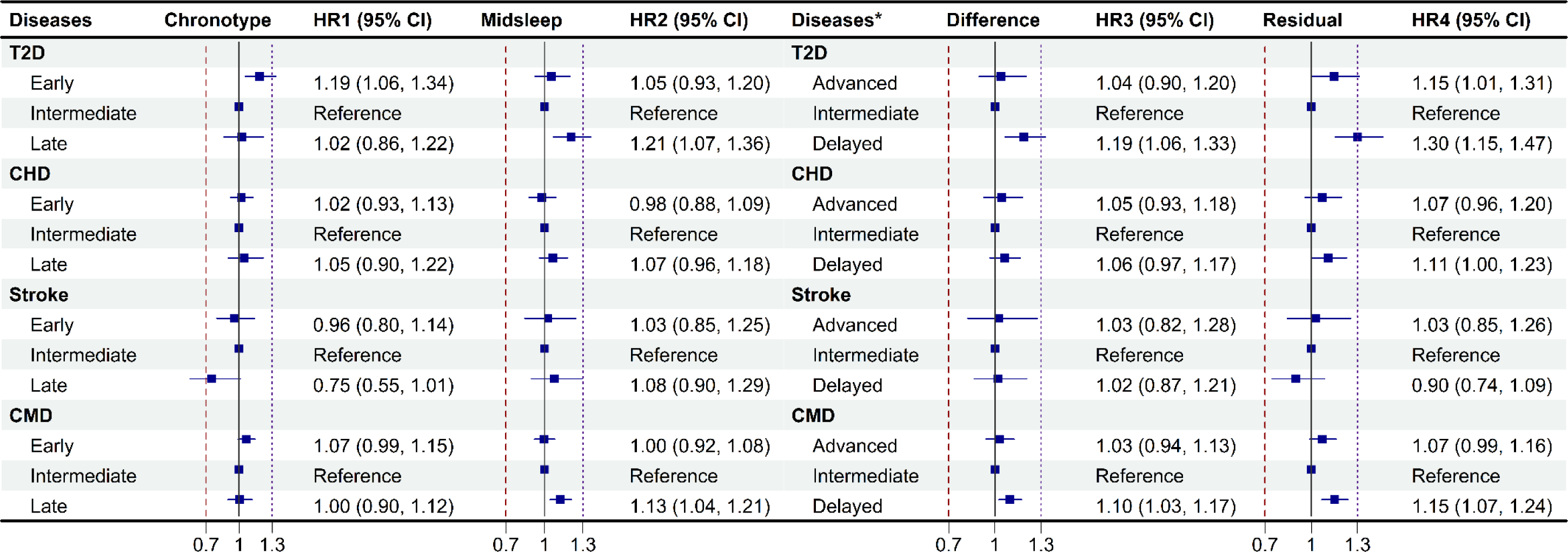
The risk of circadian preference, midsleep, and circadian misalignment on cardiometabolic outcomes. Chronotype indicated circadian preference of participants. Difference was calculated by subtracting chronotype group (early, intermediate, and late) from midsleep group (early, intermediate, and late), while Residual was calculated by the residuals of midsleep (continuous variable) on chronotype. Models were adjusted for age, sex, region, social economic status, employment, education, ethnic group, daily average noise level, smoking status, drinking status, tea consumption, coffee consumption, moderate to vigorous physic activity, and healthy sleep quality score (chronotype was omitted from healthy sleep quality score when analyzing the association between chronotype and cardiometabolic outcomes). Chronotype and midsleep were also mutually adjusted. *The category of circadian difference and residuals are different from chronotype and midsleep. Participants can be categorized into early, intermediate, and late group by chronotype and midsleep, whereas they were categorized into advanced, intermediate, and delayed sleep-wake cycle group compare to their chronotype. Abbreviation: T2D, Type 2 diabetes; CHD, coronary heart diseases; CMD, cardiometabolic diseases (incident of any of T2D, CHD, and Stroke), HR (95%CI), hazard ratio and 95% confidence limit.

**Table 2.**
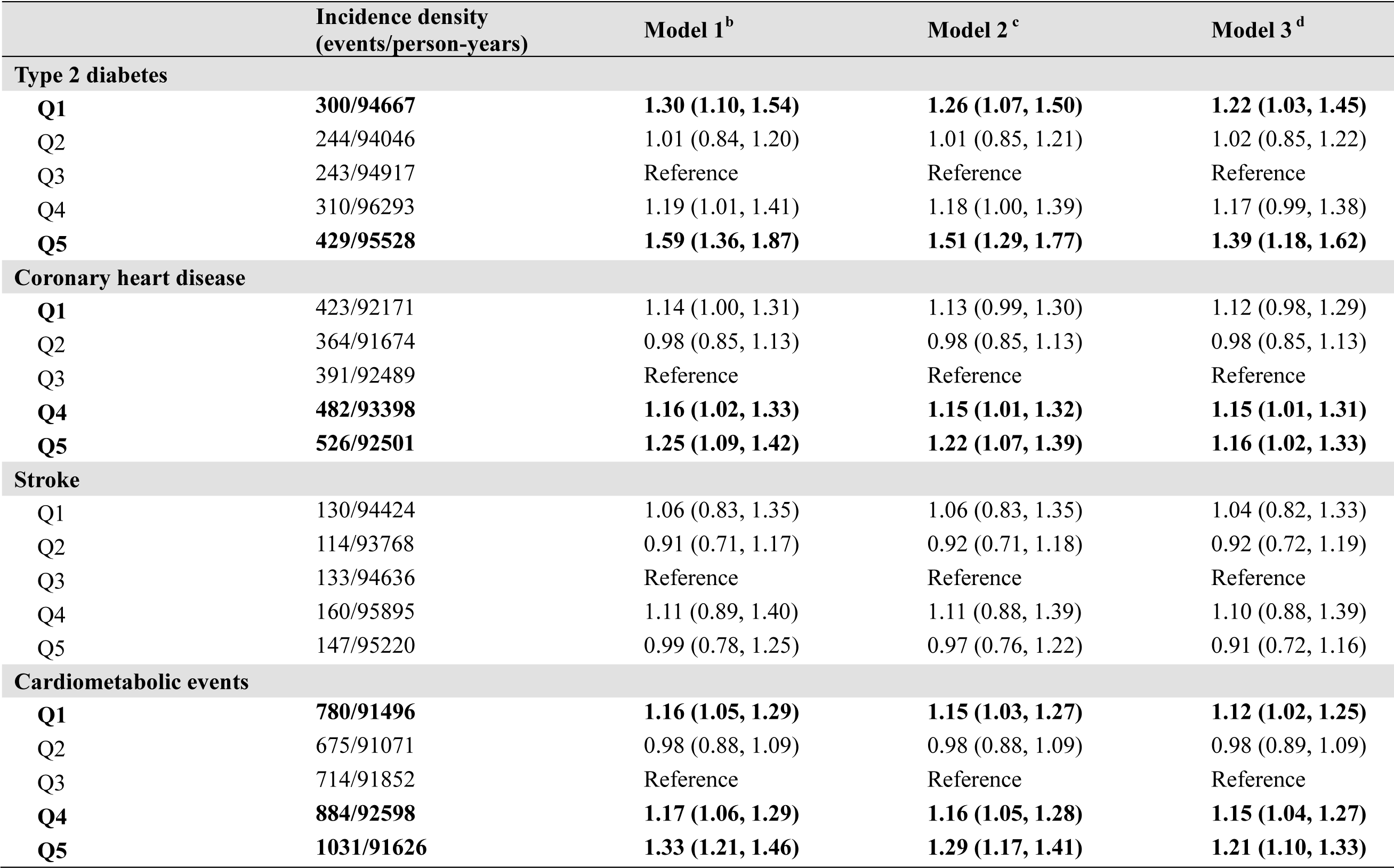
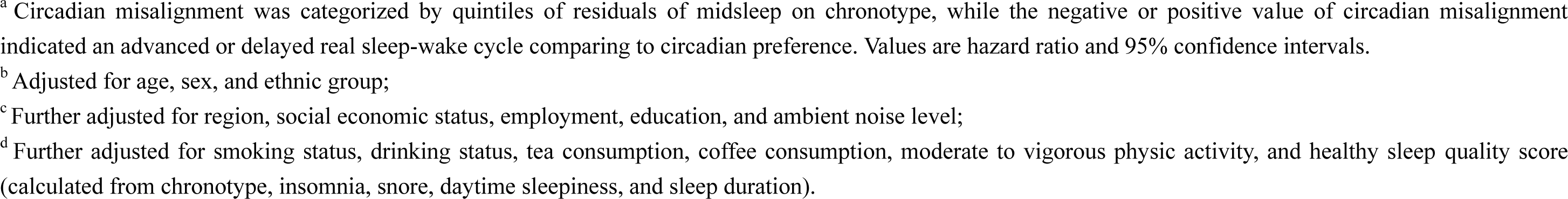
Associations between quintiles of circadian misalignment and cardiometabolic outcomes by Cox proportionate hazard models ^a^.

|  | **Incidence density**<br>**(events/person-years)** | **Model 1<sup>b</sup>** | **Model 2<sup>c</sup>** | **Model 3<sup>d</sup>** |
| --- | --- | --- | --- | --- |
| **Type 2 diabetes** |  |  |  |  |
| **Q1** | **300/94667** | **1.30 (1.10, 1.54)** | **1.26 (1.07, 1.50)** | **1.22 (1.03, 1.45)** |
| Q2 | 244/94046 | 1.01 (0.84, 1.20) | 1.01 (0.85, 1.21) | 1.02 (0.85, 1.22) |
| Q3 | 243/94917 | Reference | Reference | Reference |
| Q4 | 310/96293 | 1.19 (1.01, 1.41) | 1.18 (1.00, 1.39) | 1.17 (0.99, 1.38) |
| **Q5** | **429/95528** | **1.59 (1.36, 1.87)** | **1.51 (1.29, 1.77)** | **1.39 (1.18, 1.62)** |
| **Coronary heart disease** |  |  |  |  |
| **Q1** | **423/92171** | **1.14 (1.00, 1.31)** | **1.13 (0.99, 1.30)** | **1.12 (0.98, 1.29)** |
| Q2 | 364/91674 | 0.98 (0.85, 1.13) | 0.98 (0.85, 1.13) | 0.98 (0.85, 1.13) |
| Q3 | 391/92489 | Reference | Reference | Reference |
| **Q4** | **482/93398** | **1.16 (1.02, 1.33)** | **1.15 (1.01, 1.32)** | **1.15 (1.01, 1.31)** |
| **Q5** | **526/92501** | **1.25 (1.09, 1.42)** | **1.22 (1.07, 1.39)** | **1.16 (1.02, 1.33)** |
| **Stroke** |  |  |  |  |
| Q1 | 130/94424 | 1.06 (0.83, 1.35) | 1.06 (0.83, 1.35) | 1.04 (0.82, 1.33) |
| Q2 | 114/93768 | 0.91 (0.71, 1.17) | 0.92 (0.71, 1.18) | 0.92 (0.72, 1.19) |
| Q3 | 133/94636 | Reference | Reference | Reference |
| Q4 | 160/95895 | 1.11 (0.89, 1.40) | 1.11 (0.88, 1.39) | 1.10 (0.88, 1.39) |
| Q5 | 147/95220 | 0.99 (0.78, 1.25) | 0.97 (0.76, 1.22) | 0.91 (0.72, 1.16) |
| **Cardiometabolic events** |  |  |  |  |
| **Q1** | **780/91496** | **1.16 (1.05, 1.29)** | **1.15 (1.03, 1.27)** | **1.12 (1.02, 1.25)** |
| Q2 | 675/91071 | 0.98 (0.88, 1.09) | 0.98 (0.88, 1.09) | 0.98 (0.89, 1.09) |
| Q3 | 714/91852 | Reference | Reference | Reference |
| **Q4** | **884/92598** | **1.17 (1.06, 1.29)** | **1.16 (1.05, 1.28)** | **1.15 (1.04, 1.27)** |
| **Q5** | **1031/91626** | **1.33 (1.21, 1.46)** | **1.29 (1.17, 1.41)** | **1.21 (1.10, 1.33)** |

<sup>a</sup> Circadian misalignment was categorized by quintiles of residuals of midsleep on chronotype, while the negative or positive value of circadian misalignment indicated an advanced or delayed real sleep-wake cycle comparing to circadian preference. Values are hazard ratio and 95% confidence intervals.
<sup>b</sup> Adjusted for age, sex, and ethnic group;
<sup>c</sup> Further adjusted for region, social economic status, employment, education, and ambient noise level;
<sup>d</sup> Further adjusted for smoking status, drinking status, tea consumption, coffee consumption, moderate to vigorous physic activity, and healthy sleep quality score (calculated from chronotype, insomnia, snore, daytime sleepiness, and sleep duration).

The “U-shaped” nonlinear associations of circadian misalignment with T2D (*P_nonlinear_*<0.001), CHD (*P_nonlinear_*=0.03), and CMD (*P_nonlinear_*<0.001) were significantly revealed through RCS plots (**Figure S2**). To detect the robustness of the relationship between circadian misalignment and cardiometabolic outcomes, sensitivity analyses were conducted and yielded consistent results (**Table S4, Table S5, Figure S3**). In stratified analysis, consistent results among different ethnicity, residence, socioeconomic status, and sleep quality subgroups were displayed. Notably, the association between delayed circadian misalignment and CMDs was more prominent in women (for T2D, *P_interaction_*=0.03) or in population aged younger than 57 years, the average age (for CHD, *P_interaction_*=0.02) compared to their counterparts (**Figure S4**).

### Mediation analysis on the relationship between circadian misalignment and incident cardiometabolic events

Mediation analysis indicated that liver function, blood lipid and glucose metabolism, inflammatory factors, and anthropometric indices were associated with both circadian misalignment and CMDs, independent of demographic, socioeconomics, and lifestyle factors (**Table S6-S8**). Among them, anthropometric indices including BMI, WHR, and blood pressure introduced the largest proportion of mediation effect (44.6%) between circadian misalignment and T2D, whereas liver function, blood lipids and glucose metabolism, and inflammatory factors mediated 12.3%, 24.5%, 20.0% association between circadian misalignment and T2D, respectively **(Table 3**). As for CHD, liver function, blood lipids and glucose metabolism, inflammatory factors, and anthropometric indices introduced 8.8%, 9.1%, 13.7%, and 20.5% of mediation effects between circadian misalignment and CHD, respectively **(Table 3).**

**Table 3.**
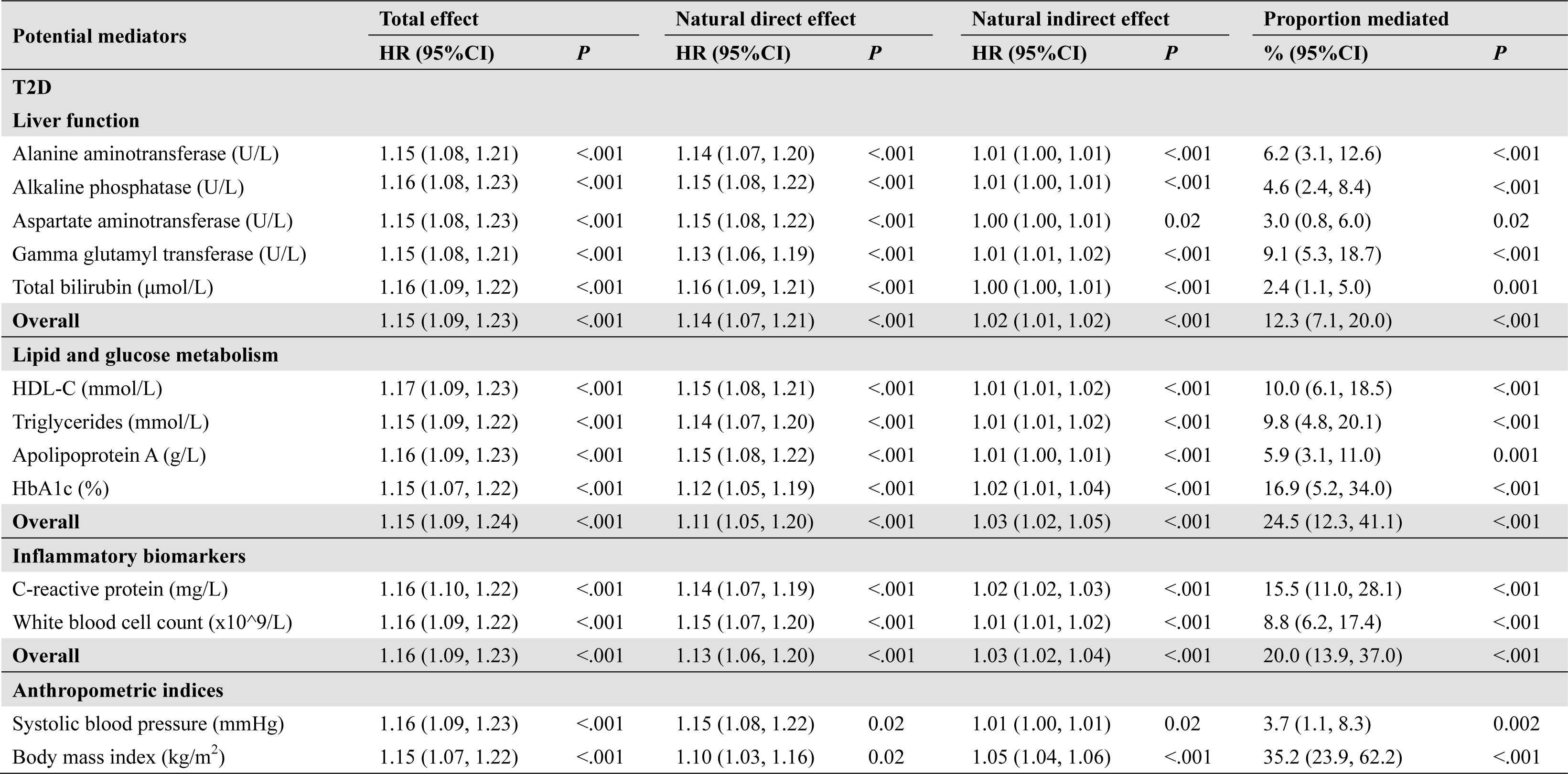
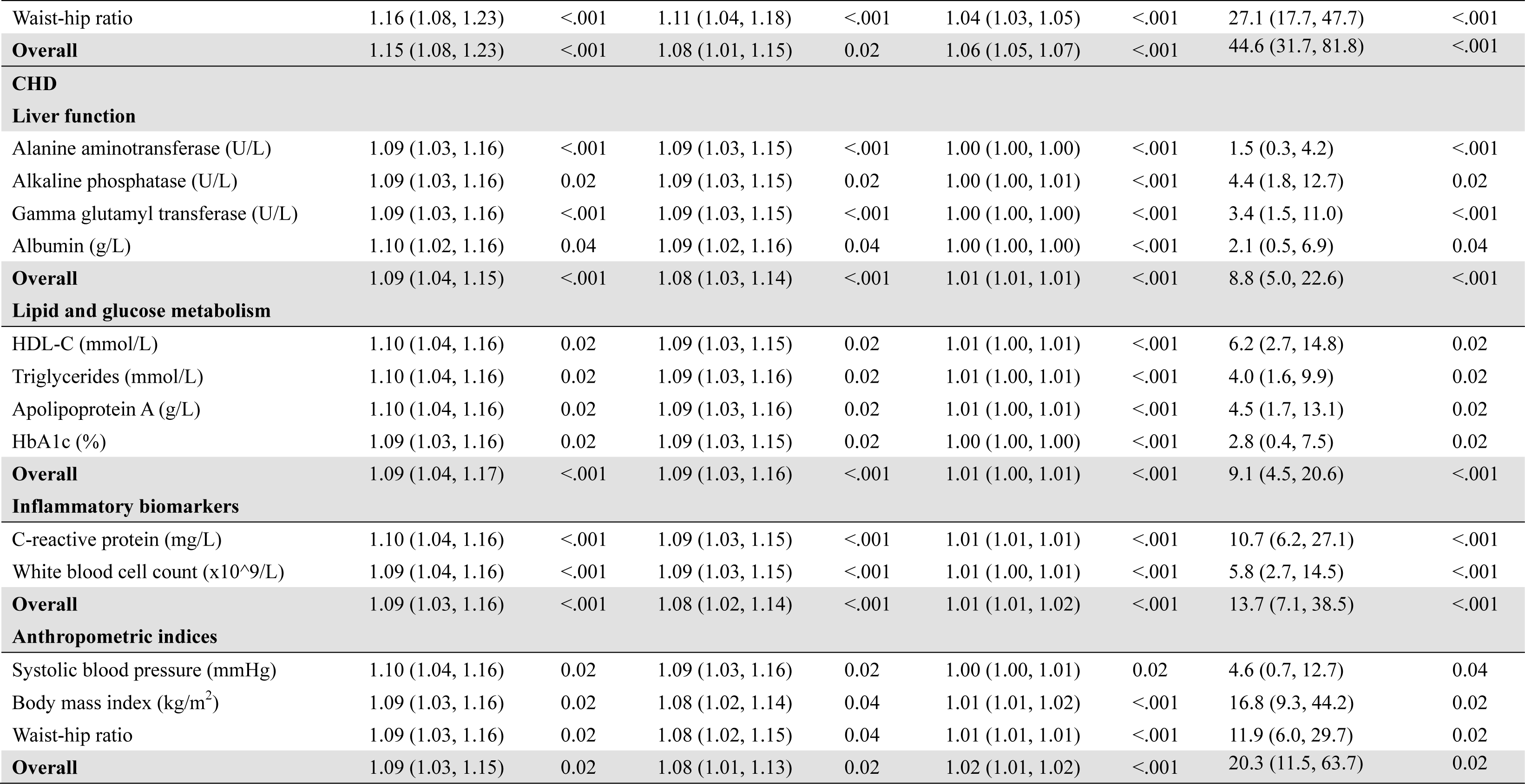
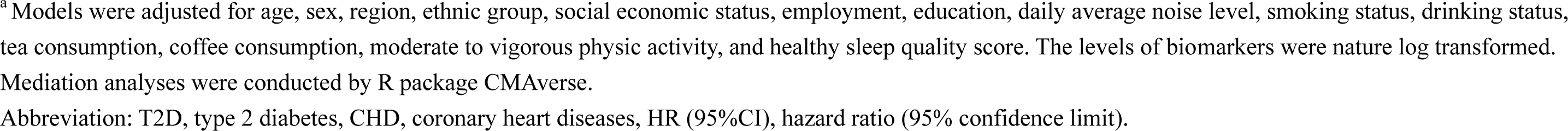
Multiple mediation models of potential mediators in the association between circadian misalignment and T2D as well as CHD^a^.

| Potential mediators | Total effect |  | Natural direct effect |  | Natural indirect effect |  | Proportion mediated |  |
| --- | --- | --- | --- | --- | --- | --- | --- | --- |
|  | HR (95%CI) | *P* | HR (95%CI) | *P* | HR (95%CI) | *P* | % (95%CI) | *P* |
| **T2D** |  |  |  |  |  |  |  |  |
| **Liver function** |  |  |  |  |  |  |  |  |
| Alanine aminotransferase (U/L) | 1.15 (1.08, 1.21) | <.001 | 1.14 (1.07, 1.20) | <.001 | 1.01 (1.00, 1.01) | <.001 | 6.2 (3.1, 12.6) | <.001 |
| Alkaline phosphatase (U/L) | 1.16 (1.08, 1.23) | <.001 | 1.15 (1.08, 1.22) | <.001 | 1.01 (1.00, 1.01) | <.001 | 4.6 (2.4, 8.4) | <.001 |
| Aspartate aminotransferase (U/L) | 1.15 (1.08, 1.23) | <.001 | 1.15 (1.08, 1.22) | <.001 | 1.00 (1.00, 1.01) | 0.02 | 3.0 (0.8, 6.0) | 0.02 |
| Gamma glutamyl transferase (U/L) | 1.15 (1.08, 1.21) | <.001 | 1.13 (1.06, 1.19) | <.001 | 1.01 (1.01, 1.02) | <.001 | 9.1 (5.3, 18.7) | <.001 |
| Total bilirubin (μmol/L) | 1.16 (1.09, 1.22) | <.001 | 1.16 (1.09, 1.21) | <.001 | 1.00 (1.00, 1.01) | <.001 | 2.4 (1.1, 5.0) | 0.001 |
| **Overall** | 1.15 (1.09, 1.23) | <.001 | 1.14 (1.07, 1.21) | <.001 | 1.02 (1.01, 1.02) | <.001 | 12.3 (7.1, 20.0) | <.001 |
| **Lipid and glucose metabolism** |  |  |  |  |  |  |  |  |
| HDL-C (mmol/L) | 1.17 (1.09, 1.23) | <.001 | 1.15 (1.08, 1.21) | <.001 | 1.01 (1.01, 1.02) | <.001 | 10.0 (6.1, 18.5) | <.001 |
| Triglycerides (mmol/L) | 1.15 (1.09, 1.22) | <.001 | 1.14 (1.07, 1.20) | <.001 | 1.01 (1.01, 1.02) | <.001 | 9.8 (4.8, 20.1) | <.001 |
| Apolipoprotein A (g/L) | 1.16 (1.09, 1.23) | <.001 | 1.15 (1.08, 1.22) | <.001 | 1.01 (1.00, 1.01) | <.001 | 5.9 (3.1, 11.0) | 0.001 |
| HbA1c (%) | 1.15 (1.07, 1.22) | <.001 | 1.12 (1.05, 1.19) | <.001 | 1.02 (1.01, 1.04) | <.001 | 16.9 (5.2, 34.0) | <.001 |
| **Overall** | 1.15 (1.09, 1.24) | <.001 | 1.11 (1.05, 1.20) | <.001 | 1.03 (1.02, 1.05) | <.001 | 24.5 (12.3, 41.1) | <.001 |
| **Inflammatory biomarkers** |  |  |  |  |  |  |  |  |
| C-reactive protein (mg/L) | 1.16 (1.10, 1.22) | <.001 | 1.14 (1.07, 1.19) | <.001 | 1.02 (1.02, 1.03) | <.001 | 15.5 (11.0, 28.1) | <.001 |
| White blood cell count (x10<sup>9</sup>/L) | 1.16 (1.09, 1.22) | <.001 | 1.15 (1.07, 1.20) | <.001 | 1.01 (1.01, 1.02) | <.001 | 8.8 (6.2, 17.4) | <.001 |
| **Overall** | 1.16 (1.09, 1.23) | <.001 | 1.13 (1.06, 1.20) | <.001 | 1.03 (1.02, 1.04) | <.001 | 20.0 (13.9, 37.0) | <.001 |
| **Anthropometric indices** |  |  |  |  |  |  |  |  |
| Systolic blood pressure (mmHg) | 1.16 (1.09, 1.23) | <.001 | 1.15 (1.08, 1.22) | 0.02 | 1.01 (1.00, 1.01) | 0.02 | 3.7 (1.1, 8.3) | 0.002 |
| Body mass index (kg/m<sup>2</sup>) | 1.15 (1.07, 1.22) | <.001 | 1.10 (1.03, 1.16) | 0.02 | 1.05 (1.04, 1.06) | <.001 | 35.2 (23.9, 62.2) | <.001 |
| Waist-hip ratio | 1.16 (1.08, 1.23) | <.001 | 1.11 (1.04, 1.18) | <.001 | 1.04 (1.03, 1.05) | <.001 | 27.1 (17.7, 47.7) | <.001 |
| **Overall** | 1.15 (1.08, 1.23) | <.001 | 1.08 (1.01, 1.15) | 0.02 | 1.06 (1.05, 1.07) | <.001 | 44.6 (31.7, 81.8) | <.001 |
| **CHD** |  |  |  |  |  |  |  |  |
| **Liver function** |  |  |  |  |  |  |  |  |
| Alanine aminotransferase (U/L) | 1.09 (1.03, 1.16) | <.001 | 1.09 (1.03, 1.15) | <.001 | 1.00 (1.00, 1.00) | <.001 | 1.5 (0.3, 4.2) | <.001 |
| Alkaline phosphatase (U/L) | 1.09 (1.03, 1.16) | 0.02 | 1.09 (1.03, 1.15) | 0.02 | 1.00 (1.00, 1.01) | <.001 | 4.4 (1.8, 12.7) | 0.02 |
| Gamma glutamyl transferase (U/L) | 1.09 (1.03, 1.16) | <.001 | 1.09 (1.03, 1.15) | <.001 | 1.00 (1.00, 1.00) | <.001 | 3.4 (1.5, 11.0) | <.001 |
| Albumin (g/L) | 1.10 (1.02, 1.16) | 0.04 | 1.09 (1.02, 1.16) | 0.04 | 1.00 (1.00, 1.00) | <.001 | 2.1 (0.5, 6.9) | 0.04 |
| **Overall** | 1.09 (1.04, 1.15) | <.001 | 1.08 (1.03, 1.14) | <.001 | 1.01 (1.01, 1.01) | <.001 | 8.8 (5.0, 22.6) | <.001 |
| **Lipid and glucose metabolism** |  |  |  |  |  |  |  |  |
| HDL-C (mmol/L) | 1.10 (1.04, 1.16) | 0.02 | 1.09 (1.03, 1.15) | 0.02 | 1.01 (1.00, 1.01) | <.001 | 6.2 (2.7, 14.8) | 0.02 |
| Triglycerides (mmol/L) | 1.10 (1.04, 1.16) | 0.02 | 1.09 (1.03, 1.16) | 0.02 | 1.01 (1.00, 1.01) | <.001 | 4.0 (1.6, 9.9) | 0.02 |
| Apolipoprotein A (g/L) | 1.10 (1.04, 1.16) | 0.02 | 1.09 (1.03, 1.16) | 0.02 | 1.01 (1.00, 1.01) | <.001 | 4.5 (1.7, 13.1) | 0.02 |
| HbA1c (%) | 1.09 (1.03, 1.16) | 0.02 | 1.09 (1.03, 1.15) | 0.02 | 1.00 (1.00, 1.00) | <.001 | 2.8 (0.4, 7.5) | 0.02 |
| **Overall** | 1.09 (1.04, 1.17) | <.001 | 1.09 (1.03, 1.16) | <.001 | 1.01 (1.00, 1.01) | <.001 | 9.1 (4.5, 20.6) | <.001 |
| **Inflammatory biomarkers** |  |  |  |  |  |  |  |  |
| C-reactive protein (mg/L) | 1.10 (1.04, 1.16) | <.001 | 1.09 (1.03, 1.15) | <.001 | 1.01 (1.01, 1.01) | <.001 | 10.7 (6.2, 27.1) | <.001 |
| White blood cell count (x10<sup>9</sup>/L) | 1.09 (1.04, 1.16) | <.001 | 1.09 (1.03, 1.15) | <.001 | 1.01 (1.00, 1.01) | <.001 | 5.8 (2.7, 14.5) | <.001 |
| **Overall** | 1.09 (1.03, 1.16) | <.001 | 1.08 (1.02, 1.14) | <.001 | 1.01 (1.01, 1.02) | <.001 | 13.7 (7.1, 38.5) | <.001 |
| **Anthropometric indices** |  |  |  |  |  |  |  |  |
| Systolic blood pressure (mmHg) | 1.10 (1.04, 1.16) | 0.02 | 1.09 (1.03, 1.16) | 0.02 | 1.00 (1.00, 1.01) | 0.02 | 4.6 (0.7, 12.7) | 0.04 |
| Body mass index (kg/m<sup>2</sup>) | 1.09 (1.03, 1.16) | 0.02 | 1.08 (1.02, 1.14) | 0.04 | 1.01 (1.01, 1.02) | <.001 | 16.8 (9.3, 44.2) | 0.02 |
| Waist-hip ratio | 1.09 (1.03, 1.16) | 0.02 | 1.08 (1.02, 1.15) | 0.04 | 1.01 (1.01, 1.01) | <.001 | 11.9 (6.0, 29.7) | 0.02 |
| **Overall** | 1.09 (1.03, 1.15) | 0.02 | 1.08 (1.01, 1.13) | 0.02 | 1.02 (1.01, 1.02) | <.001 | 20.3 (11.5, 63.7) | 0.02 |

<sup>a</sup> Models were adjusted for age, sex, region, ethnic group, social economic status, employment, education, daily average noise level, smoking status, drinking status, tea consumption, coffee consumption, moderate to vigorous physic activity, and healthy sleep quality score. The levels of biomarkers were nature log transformed. Mediation analyses were conducted by R package CMAverse.
Abbreviation: T2D, type 2 diabetes, CHD, coronary heart diseases, HR (95%CI), hazard ratio (95% confidence limit).

### Association between other circadian rhythm proxies and incident cardiometabolic events

After multiple adjustments, early chronotype [HR (95%CI): 1.19 (1.06, 1.34)] and late midsleep [1.21 (1.07, 1.36)] were both associated with increased risk of incident T2D. Meanwhile, neither chronotype nor midsleep was associated with CHD or stroke **(Figure 2)**. Notably, the significant association [HR (95%CI) 1.22 (1.03, 1.44)] between late chronotype and T2D disappeared after further adjusting for midsleep [1.08 (0.91, 1.28)].

## Discussion

In this large-scale population-based cohort study, we proposed a new metrics for circadian misalignment by calculating the discrepancy between circadian preference and accelerometer-derived actual sleep-wake cycle, and prospectively revealed a U-shaped association of circadian misalignment with T2D and CHD. Our results indicated that both advanced and delayed sleep-wake cycle in relation to individual circadian preference were associated with increased risks of CMDs, although the association was stronger in the delayed than the advanced group.

Our findings on the relationship between circadian misalignment and CMDs were consistent with previous cross-sectional studies regarding circadian disruption (social jetlag^6^, shift work^11^, or alignment between light cycle and sleep cycle^12^), although circadian misalignment in this study was measured by different metrics. Additionally, the Nurses’ Health Study also revealed similar prospective association between circadian misalignment (chronotype and shift work) and T2D^13, 23^. Unlike most studies that focusing on chronotype^9^ and shift work^7^ individually, the Nurses’ Health Study^13^ was the first large cohort study to detect circadian misalignment by examining the interaction between chronotype and shift work, revealing that an increased risk of T2D was predominantly emerged in nurses with long night shifts and an early chronotype, and was also seen in participants with long daytime work but a late chronotype. However, it remains unclear whether the mismatch between chronotype and actual sleep-wake cycle was associated with adverse health outcomes, especially in population working non-shift and less-deviated from their internal clock. Our findings by considering both circadian preference and actual sleep-wake cycle, were in line with, and further expanded these discoveries in a more quantified manner regarding circadian disruption and in both men and women population.

To date, shift work and social jetlag were the most commonly used proxies for circadian disruption in large scale population-based studies^3^. Other less-employed metrics such as composite phase deviation^10^ and delayed onset of melatonin either lack employment in large population or imposed considerable burdens on study subjetcs^14, 15^. Participants working night shift^11, 31^ is a commonly used metrics for circadian disruption, not only for its easily-collected nature but also for having the highest levels of deviation relative to internal clock^3^. Night shifts represent the most strenuous shift for most individuals, however, the association of circadian disruption on diseases for vast non-shift workers with less disturbed disruption also merits to be explored. On the other hand, since put forward in 2006, social jetlag has been widely used as a good proxy for circadian disruption, and has been associated with various adverse health outcomes^32^. It was defined by the discrepancy between social clock and biological clock while employ midpoint of sleep on work days and work-free days to respectively indicate the above two clocks. However, sleep compensation^33^ made it hard to accurately indicate biological clock by midsleep on work-free days. Moreover, it is not suitable for population of retired or without fixed work. The metrics of circadian misalignment in our analyses was defined by the discordance between self-reported circadian preference and actual sleep-wake cycle, it could be analyzed as a continuous variable to detect the U-shaped relationship between circadian misalignment and CMDs. It is also able to characterize circadian disruption on population of non-shiftwork, retired, and without fixed work.

We additionally revealed that early chronotype, rather than late chronotype, was associated with increased risk of T2D, which was inconsistent with previous evidence^23, 34^. Cross-sectional studies^35, 36^ and limited prospective cohort study^23^ revealed that late or evening chronotype were associated with higher risk of disrupted glucose homeostasis and T2D. However, to our knowledge, most existing studies^35, 36^ did not explore if the relationship between chronotype and health outcomes was independent of actual sleep-wake cycle. Our analysis indicated that after adjusting for sleep-wake cycle, the risk association of late chronotype decreased to null. Meanwhile, one large prospective analysis also revealed that participants reporting a “definite evening” chronotype were more likely to exhibit unhealthy lifestyle, which might largely explain the association with increased diabetes risk^23^. Thus, the association between late chronotype and T2D may at least partially explained by late sleep-wake cycle. Notably, a previous analysis conducted in UKB population also suggested that “definite morning” was associated with higher risk of CMDs, when classifying participants into four groups (definite morning, morning, evening, definite evening) ^37^. However, most studies^8, 13^ generally categorized the chronotype into two or three groups, making it challengeable to reveal the nonlinear relationship between chronotype and health outcomes, which are needed to be confirmed in future studies.

The potential mechanism between circadian misalignment and CMDs remains unclear. A recent compendium highlighted several diurnal and circadian mechanisms underlying cardiovascular and cerebrovascular diseases^38^: Circadian misalignment may affect immune function to promote systemic inflammation and exacerbate cardiovascular damage^39^. There is also evidence that circadian misalignment increases the risk of cardiovascular events by causing hypoxia and myocardial ischemia-reperfusion injury ^40^. Besides, molecular and physiological processes that regulate cardiovascular metabolism could also be associated with circadian misalignment through the interaction of circadian genes (*CLOCK, BMAL1, period, timeless, CRY,* etc.) and signaling pathways such as classic signal-response coupling (termed reactionary mechanisms) or temporally orchestrate metabolic pathways in preparation for predicted stimuli/stresses (termed anticipatory mechanisms) ^41^. We found that the association between circadian misalignment and CMDs may at least partially mediated by liver function, lipid metabolism, inflammatory factors, and most prominently, BMI and WHR, which may shed some light on the potential mechanism linking circadian misalignment with cardiometabolic health outcomes. Moreover, effect modification of sex on the relationship of circadian misalignment with CHD and T2D was also revealed, which may be due to the sexual dimorphism of the metabolism that contributed to sex differences in CMDs^42^. Meanwhile, age-related variations in gut microbiota and metabolic profiling^43^ may partially explain the modulatory role of age on the circadian misalignment-CMD associations. Additionally, we also found that women and older participants were more likely to have delayed circadian misalignment in our study, which was associated with increased risk of CMDs.

To our knowledge, this is the first study to examine the association between misalignment of circadian preference and actual sleep-wake cycle and cardiometabolic events. Moreover, validated accelerometers were employed to objectively measure participants’ sleep-wake cycles, whereas previous measurements of sleep-wake cycles, especially in prospective studies, were mostly based on the questionnaires^44^.

Despite the above strengths, several limitations were also worth noting. In addition to the relatively older age as well as higher socioeconomic status of UKB participants, measurement bias may generate from accelerometer derived sleep-timing, as accelerometer detected inactivity time rather than sedentary time^45^. However, good agreements (89-97%) between accelerometer-derived sleep status and polysomnography have been verified^46^, and comparing to waist-worn, the wrist-worn placement could improve capability of assessing sleep behaviors^47^. Moreover, self-reported circadian preference based on a single question from MEQ was employed. As a most widely used and easily collected circadian indicator, validity of MEQ score in relation to objectively measured circadian markers^48^, as well as that of this question to overall score^22^ has been fully investigated. Another limitation is that there is an average 5.8 years of difference between the measurement of circadian preference and real sleep-wake cycle. However, evidence has revealed circadian preference is a relatively stable characteristic for human being^5^. Besides, we included the time gap between them as a potential confounder in sensitivity analysis and also tested the interaction between the time gap and major exposures, which resulted no effect modification. A previous study in UK Biobank also indicated most of basic characteristics did not change overtime^49^. Moreover, because only one measurement on actual sleep-wake cycle was conducted, eternal time bias may happen when participants change their usual sleep-wake time. In addition, using a simple time point on midsleep to indicate sleep-wake cycle may lose detailed information on the phase and amplitude of this cycle. Nevertheless, it is an easy way to refer to sleep-wake cycle and subsequently to calculate circadian misalignment, and is appropriate for conducting circadian measurement on large-scale population studies^17^. Lastly, due to the observational nature and the possibility of residual confounding, our study could not establish causality.

In conclusion, both advanced or delayed circadian misalignment were associated with increased risk of cardiometabolic events. Population may benefit from keeping an actual sleep-wake cycle in accordance with their circadian preference. This study proposed a new metrics regarding circadian disruption, and may shed some light on the prevention and management of population with high risk of developing cardiometabolic events. Our results are needed to be confirmed by further studies that objectively measure personal circadian preference.

## Author contribution

YC, XG, and LS conceived and designed this study. YC, XX, ZZ, and HD researched data. YC, XX and LH analyzed the data. YC, TG, HY, XG and LS interpreted the results and drafted the manuscript. XG and LS supervised the study. All authors contributed to the review and revision of the manuscript and agree to be accountable for all aspects of work ensuring integrity and accuracy.

## Data Availability

This research has been conducted using the UK Biobank Resource under Application Number 96083. Data from UK Biobank is accessible to eligible researchers via applying.

http://www.ukbiobank.ac.uk

## Acknowledgement

The most important acknowledgement is to the participants in UK Biobank and the members of the UK Biobank teams, as well as to all members of team of the current analysis for their valuable help. The authors also appreciate the helpful comments on this manuscript provided by the reviewers and editors.

## Sources of funding

This study was funded by the Major Project of the Ministry of Science and Technology of China (2023YFC2506704) and the Startup grant from Fudan University (JIF201068Y and IDF201026Y).

## Disclosures

The authors declare no conflict of interest.

## Data availability

This research has been conducted using the UK Biobank Resource under Application Number 96083. Data from UK Biobank is accessible to eligible researchers via applying to www.ukbiobank.ac.uk.

## Ethic approvement

The UK Biobank was granted ethical approval from the North West Multi-center Research Ethical Committee (reference # 11/NW/0382) and research is performed in accordance with the Declaration of Helsinki.

## Notes

### Competing Interest Statement

The authors have declared no competing interest.

